# Quantification of [^11^C]ABP688 binding in human brain using cerebellum as reference region: biological interpretation and limitations

**DOI:** 10.1101/2024.02.12.24302279

**Authors:** Michele S Milella, Luciano Minuzzi, Chawki Benkelfat, Jean-Paul Soucy, Alexandre Kirlow, Esther Schirrmacher, Mark Angle, Jeroen AJ Verhaeghe, Gassan Massarweh, Andrew J Reader, Antonio Aliaga, Jose Eduardo Peixoto-Santos, Marie-Christine Guiot, Eliane Kobayashi, Pedro Rosa-Neto, Marco Leyton

## Abstract

*In vitro* data from primates provide conflicting evidence about the cerebellum’s suitability as a reference region for quantifying type 5 metabotropic glutamate receptor (mGluR5) binding parameters with positron emission tomography (PET). To address this, we first measured mGluR5 density in postmortem human cerebellum using [^3^H]ABP688 autoradiography (n=5) and immunohistochemistry (n=6). Next, *in vivo* experiments were conducted in healthy volunteers (n=6) using a high-resolution PET scanner (HRRT) to compare [^11^C]ABP688 binding potential (BP_ND_) values obtained with reference tissue methods and the two-tissue compartment model vs. metabolite-corrected arterial input function. The postmortem data showed that, relative to the hippocampus, the cerebellum had 26% less mGluR5 immunoreactivity and 94% fewer [^3^H]ABP688 binding sites. *In vivo* brain regional [^11^C]ABP688 BP_ND_ values using the cerebellum as a reference region were highly correlated with BP_ND_ values and distribution volumes derived by arterial input methods (R^2^ > 0.9). The absence of cerebellar allosteric binding sites might reflect the presence of distinct mGluR5 isoforms or conformational state. Together with our PET data, these results support the proposition that [^11^C]ABP688 BP_ND_ using cerebellum as a reference region provides accurate quantification of mGluR5 allosteric binding *in vivo*.

## 1. Introduction

Metabotropic glutamate receptors (mGluRs) modulate various aspects of glutamatergic neurotransmission. The eight known mGluRs are classified into three families according to their pharmacological properties (Kew & Kemp, 2005). mGluR5 belong to group I, which is functionally linked to the release of intracellular calcium, activation of phospholipase C, diacylglycerol, protein kinase C and inositol triphosphate (Gasparini et al., 2008).

Studies in both laboratory animals and humans provide evidence that mGluR5 are involved in dementia (Parameshwaran et al., 2008), anxiety (Spooren et al., 2000), motor (Breysse et al., 2002) and substance use disorders (Chiamulera et al., 2001; Akkus et al., 2013; Milella et al., 2014; Martinez et al., 2014; Cox et al., 2020). Moreover, converging evidence points toward a specific role for mGluR5 in molecular mechanisms of neuroprotection (Bruno et al., 2001) and synaptic plasticity (Gladding et al., 2009). Given these features, quantification of receptor availability *in vivo* is crucial for testing mGluR5 hypotheses in people with neuropsychiatric disorders and evaluating novel therapeutic targets.

In contrast to most positron emission tomography (PET) receptor probes, (3-(6-methyl-pyridin-2-ylethynyl)-cyclohex-2-enone-O-11C-methyl-oxime ([^11^C]ABP688) binds with high selectivity and specificity to a transmembrane allosteric site instead of orthosteric binding sites. The *in vivo* pharmacology of imaging agents specific to allosteric binding sites is a relatively new frontier in neuroreceptor imaging since the binding pockets for allosteric activators and inhibitors are located in the membrane compartment and do not compete directly with the transmitter (Ametamey et al., 2007). Moreover, it remains unclear whether allosteric binding sites in the nervous system are exclusively available in a single conformational state.

As a PET ligand, [^11^C]ABP688 shows a favorable chemical and pharmacokinetic profile including rapid brain uptake, high mGluR5 affinity, fast kinetics, and absence of brain permeable radiolabeled metabolites (Hintermann et al., 2007). A consistent anatomical distribution pattern is seen when comparing *in vitro* and *in vivo* estimates of mGluR5 specific binding in the rodent brain (Shigemoto et al., 1993; Elmenhorst et al., 2010).

Most PET [^11^C]ABP688 studies have relied on a single bolus paradigm of tracer administration. A two-tissue compartmental model (2TCM) using metabolite-corrected plasma input function has been successfully employed to quantify mGluR5 availability in humans (Ametamey et al., 2007; Treyer et al., 2007; DeLorenzo et al., 2011a), baboons (DeLorenzo et al., 2011b; Miyake et al., 2011) and rats (Wyss et al., 2007; Helmenhorst et al., 2010).

Non-invasive quantification of [^11^C]ABP688 binding parameters (i.e., binding potentials; BP_ND_) without arterial blood sampling has been validated in non-human species, using the cerebellum as reference region (Helmenhorst et al., 2010; DeLorenzo et al., 2011b). These methods are based on extremely low mGluR5 concentrations in the rodent cerebellum (Ametamey et al., 2006; Treyer et al., 2007). Using quantification of *in vivo-in vitro* binding parameters in the rodent brain, we have found that the saturable [^3^H]ABP688 cerebellar binding sites found *in vitro* were not sensitive to competitive blocking with MPEP previously measured *in vivo* with [^11^C]ABP688 and PET (Helmenhorst et al., 2010). However, these results cannot necessarily be translated to human studies since Patel and colleagues (Patel et al., 2007) suggest that the presence of cerebellar mGluR5 immunoreactivity and binding site availability in humans might be higher than in rodents.

Given the conflicting evidence, our goal was to conduct *in vivo* and *in vitro* experiments to test whether the cerebellum can be considered a reference region suitable for [^11^C]ABP688 quantification in humans. To that end, we performed cerebellar and hippocampal semi-quantitative mGluR5 immunohistochemistry and [^3^H]ABP688 quantitative autoradiography to characterize binding in regions previously described as harboring high and low mGluR5 densities. Additionally, we established correlations between binding parameters obtained with PET metabolite-corrected arterial input function (AIF) and the cerebellum as a reference region. Due to safety issues and Health Canada regulations, displacement studies with co-injection of large doses of [^11^C]ABP688 competitors cannot be conducted in living human individuals. Circumventing arterial cannulation would greatly simplify the procedure and limit discomfort during testing.

## 2. Methods

### 2.1. Postmortem brain specimens and Ethics

Specimens were obtained from the Brain Bank from the Douglas Mental Health University Institute (Montreal, Canada). All *in vitro* experiments were carried out in accordance with the guidelines provided by the Douglas Brain Bank research board, approved by the Research and Ethics Board of the Douglas Research Institute, McGill University. Specimens utilized in this study were re-evaluated by a neuropathologist (MCG) to confirm the absence of brain pathology that could confound the results (i.e. cerebrovascular disease or neurodegenerative processes). Presence of ante mortem history of DSM-IV psychiatric conditions or neurological disorders was excluded based on both interview with family members and/or treating professionals, and on medical records.

### 2.2. Radiochemistry

*In vitro study:* [^3^H]ABP688 specific activity (2738 GBq/mmol) was purchased from Amersham/GE Healthcare Biosciences (Little Chalfont, Buckinghamshire, UK). [^3^H]ABP688 was synthesized from desmethyl-ABP688 cis/trans (1:8) produced by ABX (Radeberg, Germany), by reacting the sodium salt of desmethyl-ABP688 in anhydrous dimethyl-sulfoxide with [^3^H]methyl iodide at 90°C for 5 minutes.

*In vivo study:* Desmethyl-ABP688 cis/trans (1:8) was produced by ABX (Radeberg, Germany). [^11^C]ABP688 was synthesized by reacting the sodium salt of desmethyl-ABP688 in anhydrous dimethyl-sulfoxide with [^11^C]methyl iodide at 90°C for 5 minutes. The product was purified by semipreparative high-performance liquid chromatography (Waters, µBondapak, C18; mobile phase, acetonitrile: 0.1% phosphoric acid (30:70); flow rate, 2mL/min), and the retention time was 10 minutes. After removal of the high-performance liquid chromatography solvent by evaporation, the product was formulated using 9 mL of phosphate buffer and 1 mL EtOH. TLC: CH2Cl2:MeOH:AOAc (7:2:1). The radiochemical purity was > 99%. The total time required for the synthesis of [^11^C]ABP688 was 30 minutes from the end of bombardment.

### 2.3. [^3^H]ABP688 Quantitative Autoradiography

Human frozen brain slices (n=5, demographics in Table 1 of Supplementary Information) corresponding to the cerebellum and hippocampus regions were studied. Hippocampal CA1 region was selected for normalized quantification across subjects. Tissues were cryosectioned at 20 µm at -15°C (HM 500M, Microm International) and thaw-mounted on poly-L-lysine pre-coated microscope slides. Brain sections were dried at room temperature for one hour, and then stored in a freezer at -80°C until use. The *in vitro* [^3^H]ABP688 binding study was performed according to Hintermann et al. (2007) with some modifications. Briefly, slides were warmed up to room temperature and pre-incubated for 20 min in buffer containing 30 mmol N_2_ HEPES, 110 nmol NaCl, 5 mmol KCl, 2.5 mmol CaCl_2_ and 1.2 mmol MgCl_2_ (pH 7.4). A [^3^H]ABP688 saturation binding study was performed using concentrations of 8, 4, 2, 1, 0.5, 0.25 and 0.125 nM in the same buffer for 60 min at room temperature. Non-specific binding was determined with the addition of the selective, non-competitive mGluR5 antagonist 2-methyl-6- (phenylethynyl)-pyridine (MPEP, 10 µmol/L) in adjacent sections. After the incubation, slides were washed (3 × 5 minutes) in cold buffer, dipped in ice-cold distilled water and rapidly dried under a stream of cool air. After drying, tissues were fixed, desiccated by exposure to paraformaldehyde powder *in vacuo* overnight (Liberatore et al., 1999) and exposed along with [^3^H] microscales (GE Healthcare, UK) to tritium-sensitive radioluminographic imaging plates (BAS-TR, Fuji-Film, Japan) for five days. After exposition, imaging plates (BAS-TR2025, Fuji-Film) were scanned using BAS 5000 (Fuji-Film). Imaging plates were analyzed using the software ImageGauge 4.0 (FujiFilm). Specific radioactivity was calibrated using the [^3^H] microscales and measured in regions of interest (as described below). Analysis of the saturation binding data was calculated by fitting a one site binding model to the specific binding data using the GraphPad Prism 4 Software (GraphPad Software, Inc, San Diego, USA). Values for the maximum receptors density (B_max_) and dissociation constant (K_D_) are expressed in pmol/mg of wet tissue and nmol of ligand, respectively. All statistical analyses were performed using R Statistical (version 2.10.0) and GraphPad Prism 4 software.

**Table 1.** Average kinetic parameters and total distribution volumes obtained with metabolite-corrected plasma input function.

| ROI | 2TCM | | | | $V_T$ (mL/cm<sup>3</sup>) | GA Logan<br>$V_T$ (mL/cm<sup>3</sup>) |
| --- | --- | --- | --- | --- | --- | --- |
| | $K_1$ (mL/cm<sup>3</sup>/min) | $k_2$ (min<sup>-1</sup>) | $k_3$ (min<sup>-1</sup>) | $k_4$ (min<sup>-1</sup>) | | |
| cerebellum | 0.15 ± 0.03 | 0.33 ± 0.14 | 0.08 ± 0.02 | 0.03 ± 0.01 | 2.01 ± 0.48 | 1.90 ± 0.48 |
| ACC | 0.14 ± 0.02 | 0.15 ± 0.05 | 0.11 ± 0.06 | 0.05 ± 0.02 | 3.60 ± 0.59 | 3.43 ± 0.55 |
| caudate | 0.15 ± 0.03 | 0.17 ± 0.06 | 0.12 ± 0.06 | 0.04 ± 0.02 | 3.53 ± 0.99 | 3.39 ± 0.96 |
| hippocampus | 0.14 ± 0.03 | 0.22 ± 0.13 | 0.17 ± 0.10 | 0.05 ± 0.01 | 3.17 ± 0.58 | 3.04 ± 0.54 |
| PFC | 0.15 ± 0.03 | 0.20 ± 0.11 | 0.13 ± 0.07 | 0.04 ± 0.02 | 3.69 ± 0.74 | 3.58 ± 0.70 |
| putamen | 0.18 ± 0.02 | 0.22 ± 0.06 | 0.10 ± 0.03 | 0.03 ± 0.01 | 3.46 ± 0.64 | 3.39 ± 0.69 |

Values are mean ± SD ( $n = 6$ ); 2TCM, two-tissue compartment model; GA, graphical analysis; $V_T$ , total distribution volume; ACC, anterior cingulate cortex; PFC, prefrontal cortex

### 2.4. Tissue mGlu5 Receptor Immunohistochemistry

Formalin fixed human cerebellum and hippocampus (n=6; demographics in Table 1 of Supplementary Information) were studied. Specimens were paraffin-embedded and cut at 3 µm for mGluR5 immunohistochemistry, performed in a BenchMark XT (Ventana Medical System, Tucson, Arizona), following manufacturer’s protocols. We developed an automatic immunohistochemistry pipeline to automatically process all slides utilized in this study, which minimizes variability across slides due to operator interaction in individual slides. Slides were incubated with a rabbit primary antibody anti-rat mGluR5 (AB5675; Millipore) and diluted at 1:100 in blocking medium for 32 minutes at 37^°^C. The polyclonal antibody was generated by an in-frame insertion of 21 amino acids close to the C-terminus of the receptor. Revelation was performed with Ventana Ultraview DAB detection kit (Ventana Medical System), followed by counterstaining. All slides underwent quality control.

### 2.5. Immunohistochemistry analysis

Digital images from stained sections were obtained in an Aperio ScanScope AT (Aperio Technologies, Vista, California) at a 20× magnification. Quantification of strong positive areas was performed with ImageScope software (Aperio Technologies, Vista, California), using its positive pixel count algorithm. Regions of interest were drawn in different regions of the sections containing the cerebellum and hippocampus, and the number of DAB positive pixels was quantified, averaged and normalized as 100. Immunoreactivity was classified as weak (<100), moderate (between 100 and 175) and strong (>175). The final results are shown as percentage of immunopositive pixels (number of immunopositive pixels/number of immunopositive pixels + number of immunonegative pixels).

### 2.6. In vivo experiments

The study was carried out in accordance with the Declaration of Helsinki and was approved by the Research and Ethics Board of the Montreal Neurological Institute/McGill University. Written informed consent with details of the experimental procedures and approved by our institution Research Ethics Board was obtained from all subjects before scan sessions.

### 2.7. Subjects

Six healthy volunteers (2 women; 21-38 yo; mean age ± SD, 24.5 ± 6.6 y) participated in the [^11^C]ABP688 PET study, recruited from the community through online advertisement. After a pre-screening phone interview, potential candidates underwent a face-to-face interview with the investigator (MSM), with special attention to personal and family history of neurological and psychiatric disorders using a semi-structured clinical interview for DSM IV (SCID). For each candidate, a medical evaluation, electrocardiogram, routine blood test (CBC, electrolytes, thyroid, cholesterol, liver and renal workup) and urine toxicology screening were performed to assess physical health status. Volunteers were excluded in the presence of a current Axis I psychopathology or neurological disorder, a somatic disorder, and presence and/or history of first-degree relatives with neurological or psychiatric disorder, including substance use disorders. Before a PET scan session began, all participants tested negative on a urine drug screen (Triage Panel for Drugs of Abuse, Biosite Diagnostics, San Diego, California) and women were scanned during the follicular phase and tested negative on a urine pregnancy test.

### 2.8. Positron Emission Tomography Procedures

For [^11^C]ABP688 injection, a cannula was inserted in the right arm antecubital vein. A saline solution of 362.6 ± 43.3 MBq of [^11^C]ABP688 was then administered as a 1-minute intravenous slow bolus injection, and emission scanning started concurrently with the start of the bolus injection. Participants were scanned in supine position with a head-fixation device to minimize head movement during data acquisition. PET imaging was performed with a Siemens High Resolution Research Tomograph (HRRT+) scanner (CTI/Siemens, Knoxville, Tennessee), which combines high spatial image resolution with high sensitivity (de Jong et al., 2007). It consists of 8 detector heads arranged in an octagon. A detector head comprises 117 detector blocks, each cut into 8 × 8 crystal elements. Each block consists of 2 lutetium oxyorthosilicate/lutetium-yttrium oxyorthosilicate (LSO/LYSO) crystal layers to achieve photon detection with depth-of-interaction information. The spatial resolution range of the scanner is between 2.3 and 3.4 mm full width at half maximum. A 7-min transmission scan was acquired before the emission scan for attenuation correction. The PET emission data were acquired in list-mode format and binned into 26 time frames. For every time frame, fully 3D sets of sinograms were generated from the list-mode data (2209 sinograms, span 9, with 256 radial bins and 288 azimuthal angle samples). A time-series of 26 3D images (frames) were then reconstructed from these sinograms, each 3D image being composed of 256 × 256 × 207 cubic voxels (voxel side-length of 1.21875 mm), using an expectation maximization image reconstruction algorithm with an ordinary Poisson model of the acquired PET data (see Reader et al., 2006). The reconstruction included full accounting for the normalization, attenuation, and time-dependent scatter and randoms. Subject head-motion was corrected for using a data-based motion estimation and correction method (Costes et al., 2009).

### 2.9. Arterial blood sampling and metabolite analysis

For AIF, a catheter was inserted in the radial artery by a certified anesthesiologist (MA). During the scan, 19 sequential 1 ml blood samples were manually collected. Each sample was drawn by releasing the blockade of the catheter after discarding the dead volume in the 1 mm diameter tube of a three-way stopcock. Catheters were flushed with heparinized saline solution after each sampling. For the first minute, samples were taken every 10 seconds, with increasing intervals afterwards until the end of the scan (2x30s, 3x1min, 1x2min, 1x3, 5x10min). After separation of an aliquot of whole blood, each sample was centrifuged to obtain 200 μL plasma aliquots. The concentration of radioactivity in whole blood and plasma aliquots was then measured in a gamma counter cross-calibrated with the PET system. Six additional samples of 5 mL were collected for metabolites determination at 1, 2, 5, 15, 30, 60 minutes. The percentage of unchanged radioligand in the plasma was determined as described earlier (Wyss et al., 2007). Briefly, lipophilic parent compound is retained by the solid phase of C18 cartridges (Sep-Pak Plus; Waters Corporation, Milford, MA, USA). The cartridges were prewashed with 5 mL of water and flushed with the plasma sample diluted in 2.5 mL of water. Hydrophilic compounds were then eluted with 5 mL of water, and the cartridge was flushed with 10 mL of air to clear from fluid. The cartridge and a 1 mL fraction of the flushing fluid were then measured in the well counter. Previous test revealed that the retention of the parent compound in solvent to the cartridge (recovery factor) was between 91% and 96%. Therefore, the time course of the fraction of parent compound was normalized to the first sample and was fitted by nonlinear regression analyses (*f*(t) = 1/(1 + *a* × *t*^*b*)^*c* + *d*). This fit was subsequently used to generate metabolite-corrected plasma curves.

### 2.10 Image Analysis

For anatomical co-registration, a high-resolution (1 mm) T1-weighted magnetic resonance image (MRI) acquisition was obtained for all subjects on a 1.5 T Siemens Sonata scanner, using gradient echo pulse sequence (repetition time = 9.7 msec, echo time = 4 msec, flip angle = 12°, field of view = 250 and matrix 256 × 256). Each subject’s MRI volume was manually co-registered with the 4D reconstructed PET image using the MINC tools package (McConnell Brain Imaging Center, McGill University, Montreal, Canada; http://www.bic.mni.mcgill.ca/ServicesSoftware/MINC). Regions of interest (ROIs) were manually drawn on the MRI co-registered with PET images in native space: cerebellar cortex, caudate, putamen, hippocampus, prefrontal cortex (PFC) and anterior cingulate cortex (ACC). Corresponding time-activity curves (TACs) were generated with regional activity concentration calculated for each frame, corrected for decay and plotted versus time.

### 2.11. Kinetic Models

The outcome parameters measured are the equilibrium total distribution volume (V_T_) and binding potential (BP_ND_), that are directly proportional to receptor density (*B_max_*) (Innis et al., 2007). The V_T_ is composed by the specific distribution volume (V_S_, equal to f_P_B_avail_/K_D_) and the distribution volume of free and nonspecifically bound ligand (V_ND_, which is assumed to be equivalent to the volume of distribution of the reference region). While V_T_ is dependent on blood sampling, BP_ND_ refers to a region void or with negligible specific binding and is therefore independent from AIF. The gray matter of the cerebellum was chosen as reference region throughout all the modeling. Kinetic analysis was performed using the ifit software package (Kjærgaard et al., 2021).

*V_T_ estimation.* A 2TCM with four unconstrained kinetic constants was applied to analyze the data. 2TCM has been previously shown to provide superior fits than the one-tissue compartment model for [^11^C]ABP688 in all brain regions, cerebellum included (Treyer et al., 2007). Vascular contribution to tissue TACs (V_b_) was added as an additional unfixed parameter in the model. The rate constants for each ROI were determined from the TACs by non-linear curve fitting in the least-squares sense. V_T_ was defined as (K_1_/k_2_)(1 + k_3_/k_4_). To test for higher stability of the fits, a constrained 2TCM was also applied. The ratio K_1_/k_2_ in target ROIs was fixed to the value (∼10%) estimated in the reference region, under the assumption that it is similar in different brain regions.

V_T_ was also estimated using the Logan’s graphical analysis (GA) (Logan et al., 1990). In this simplified approach, which uses plasma as input function, V_T_ corresponds to the slope of the linear portion of the resulting plot for each ROI. V_b_ contribution was fixed to the values derived in the 2TCM analysis. Time of linearization was set to t* = 10 minutes.

*Calculation of BP_ND_* was determined by the following methods: (1) distribution volume ratio (DVR) derived from the 2TCM (V_T_/V_ND_−1); (2) simplified reference tissue method (SRTM) (Lammertsma & Hume, 1996) in which the reference region TAC is used as input function to interpret TACs of target regions through non-linear curve fitting procedure; (3) non-invasive Logan’s GA (NIGA) with a t* = 10 minutes (Logan et al., 1996). This modified approach uses the reference region as input function and allows for the estimation of the DVR. BP_ND_ is calculated as DVR-1. Selection of t* was based on visual inspection of residual plot.

### 2.12. Statistics

All values are expressed as mean ± SD. For the kinetic study, the adequacy of the fitting for each model was judged according to the Akaike information criterion (AIC), with lower AIC values indicative of a better fit. Pearson’s correlation coefficient and linear regressions were performed using SPSS (IBM SPSS Statistics, version 20.0.0) as well as ANOVA two-way analysis when needed.

## 3. Results

### 3.1. In vitro experiments

#### 3.1.1. Autoradiography and saturation binding analysis

The mean B_max_ values were 1.31 ± 0.1 and 0.08 ± 0.01 pmol/mg of tissue for CA1 region and cerebellum (t-test, p<0.001). Figure 1A shows representative total binding in the two brain areas. There was no difference in the maximum density of mGluR5 (B*_max_*) in the molecular and granular layers of the cerebellum. Cerebellar K_D_ was also significantly lower than hippocampal K_D_ (0.4 ± 0.02 vs 1.5 ± 0.2 nmol/L, t-test, p<0.05). Hippocampus/cerebellum B_max_ ratio of 15:1 was observed. Figure 1B shows representative nonspecific binding in the presence of the competitor. MPEP (2 nmol/L) reduced [^3^H]ABP688 total binding by >75% in the cerebellum (on the left) and >95% in the hippocampus (on the right).

**Figure 1.**
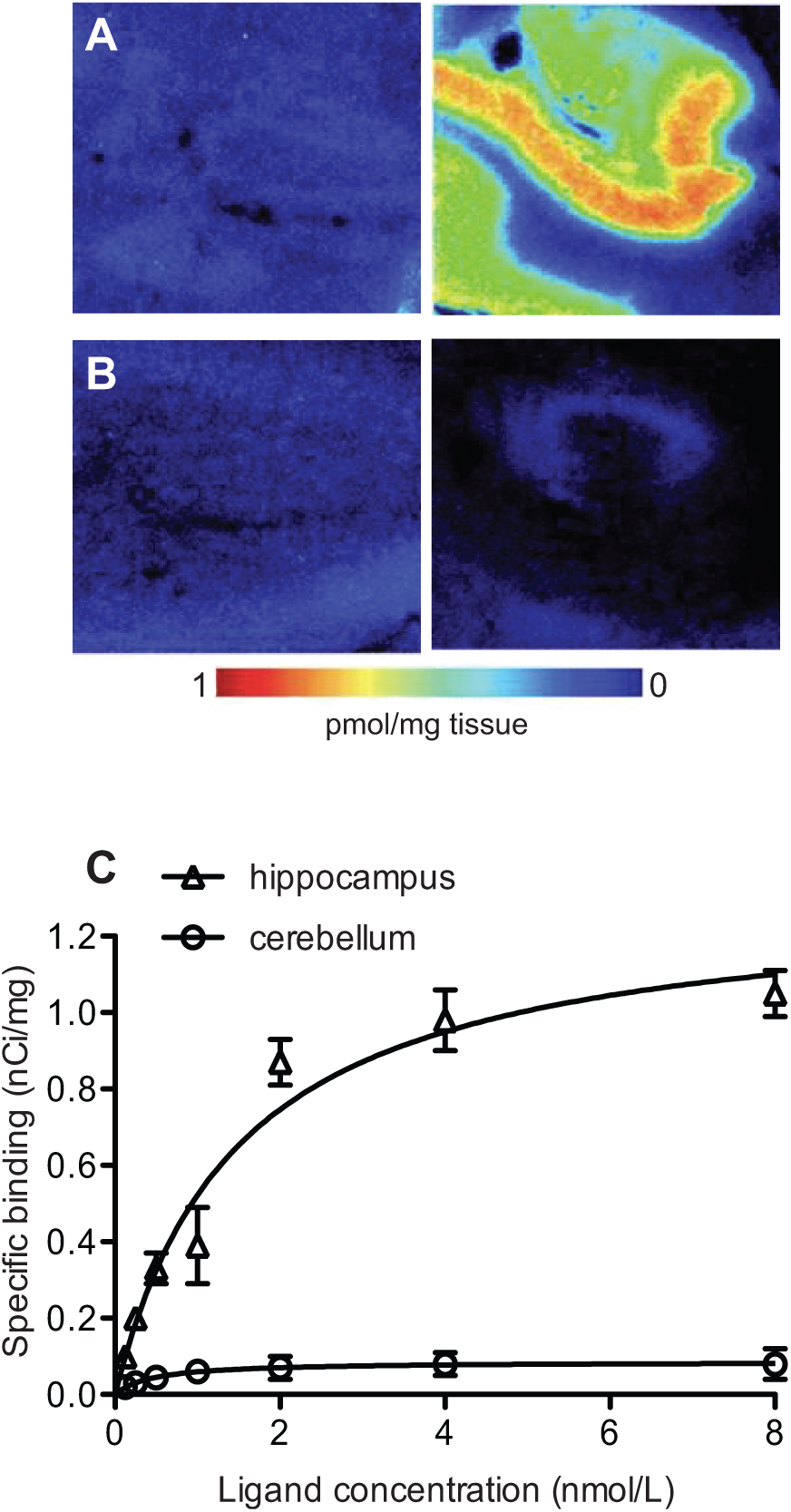
[^3^H]ABP688 binding and the effect of blockade with MPEP in the autoradiography study in human brain sections. Representative autoradiograms showing (A) the total binding of 2nmol/L [^3^H]ABP688 in the cerebellum (left) and hippocampus (right), and (B) the nonspecific binding in the presence of the antagonist MPEP, in the cerebellum (left) and hippocampus (right). The scale shows density of mGluR5 in pmol/mg of tissue. (C) Saturation binding experiment with [^3^H]ABP688. Displayed are the nonlinear regression fit curves for the CA1 region of the hippocampus (high-binding region) and the cerebellum (reference region).

#### 3.1.2. Immunohistochemistry

mGluR5 immunoreactivity was definitively positive in the human cerebellum (Figure 2A & 2B). Moderate immunoreactivity was homogeneously observed throughout the molecular and granular layers of the cerebellar cortex. Purkinje cell layer was strongly immunoreactive. Neurons of the dentate nucleus showed only moderate immunoreactivity. No significant staining was observed in the white matter. In the hippocampus (Figure 2C & 2D) stronger mGluR5 staining was observed in pyramidal neurons. mGluR5 staining was less pronounced in granular cells. The quantification of positive areas revealed a higher percentage of mGluR5 immunopositive area in the hippocampus than in the cerebellum (Figure 2E, hippocampus = 58.7 ± 19.8%, cerebellum = 42.2 ± 21.6%; t-test, p<0.001).

**Figure 2.**
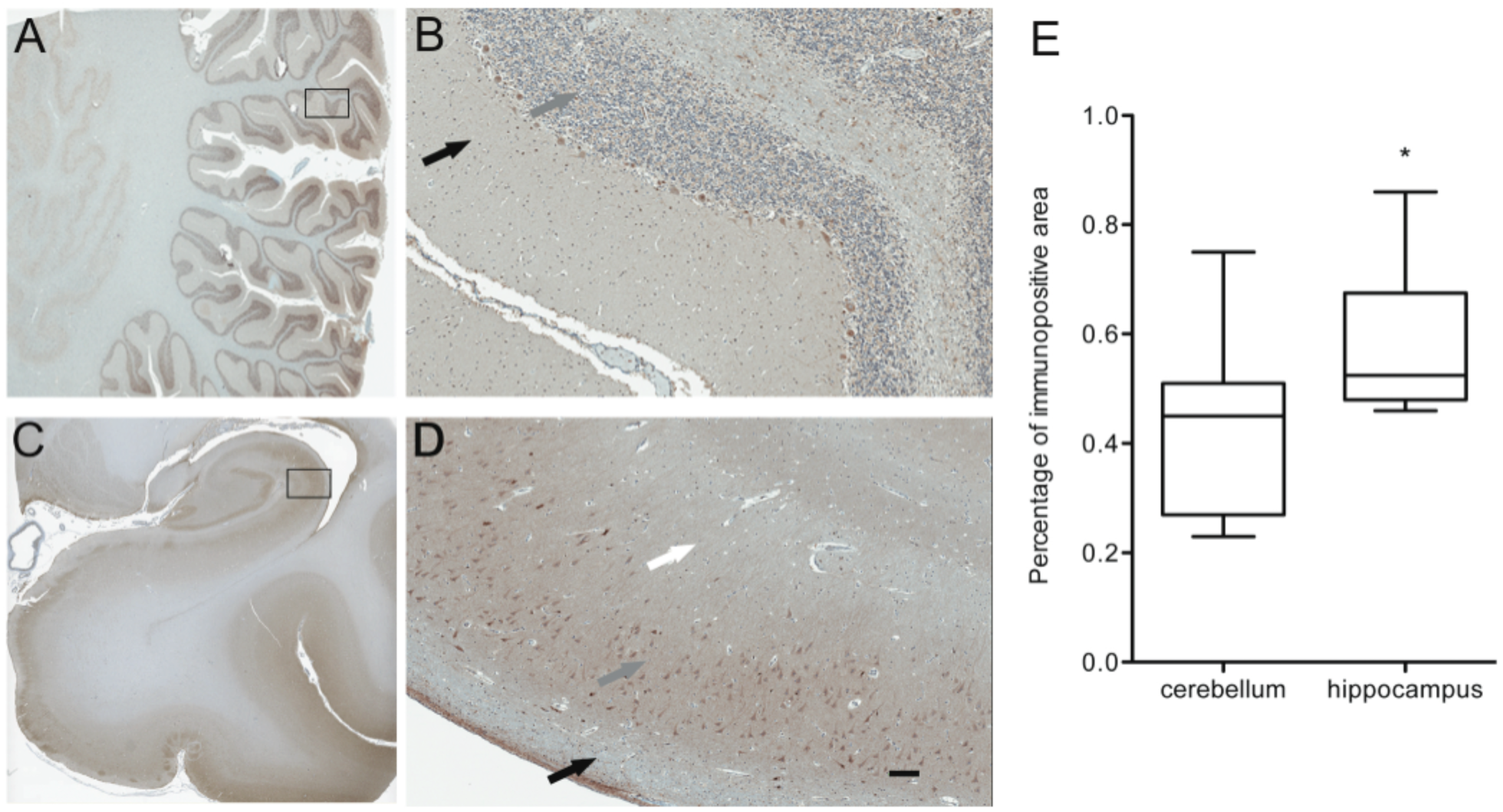
Distribution of mGluR5 in human brain sections of cerebellum (A,B) and hippocampus (C,D) at immunohistochemistry. In the cerebellum, moderate staining is observed in the molecular layer and nuclear layer (black and grey arrow in B, respectively). In CA region of the hippocampus, strong staining is observed in the soma and neuropil of the pyramidal layer (grey arrow in D). Lower intensity of staining is observed in the dendritic branches in the stratum radiatum (white arrow in D) and in the stratum oriens (black arrow in D). The bar in D indicates 100 micrometers. (E) Quantification of mGluR5-immunopositive area in cerebellum and hippocampus. The boxplots represent the lower and upper quartile and the median of the distribution, the whiskers represent the lowest and highest value found. * p<0.001.

### 3.2. In vivo experiments

3.2.1. Regional uptake and AIF *in vivo* PET study

Figure 3 shows the curves for the total concentration of radioactivity in the plasma, the metabolite-corrected plasma activity and the fraction of parent compound as a function of time. After injection, [^11^C]ABP688 activity in the arterial plasma reached a peak at 50-70 seconds. [^11^C]ABP688 was rapidly metabolized, with metabolites accounting for approximately 50% of the radioactivity in the plasma after 5 minutes (Figure 3B). The fraction of non-metabolized parent compound decreased at a slower rate thereafter, to 0.22 ± 0.1, 0.14 ± 0.8 and 0.08 ± 0.06 at 15, 30 and 60 minutes respectively. The whole blood/plasma concentration ratio was 0.61 ± 0.12 at peak and 0.80 ± 0.12 at 60 minutes after injection. Mean specific activity of the radioligand was 0.25 ± 0.14 mCi/nmol, with mean injected mass of 0.20 ± 0.11 μg/kg of body weight. The radioligand injected was rapidly distributed in the brain, as shown in the TACs in Figure 3C. Radioactivity peaked after 1.7 minutes and the concentration was highest in the mGluR5-rich regions, such as putamen and caudate. All other areas showed comparable levels of peak estimates, although, as expected, cerebellum presented the fastest washout of radioactivity (more than halved at 20 minutes post-injection). [^11^C]ABP688 plasma clearance was 88.7 ± 24.3 L/h, calculated as the area under the curve and dose injected ratio.

**Figure 3.**
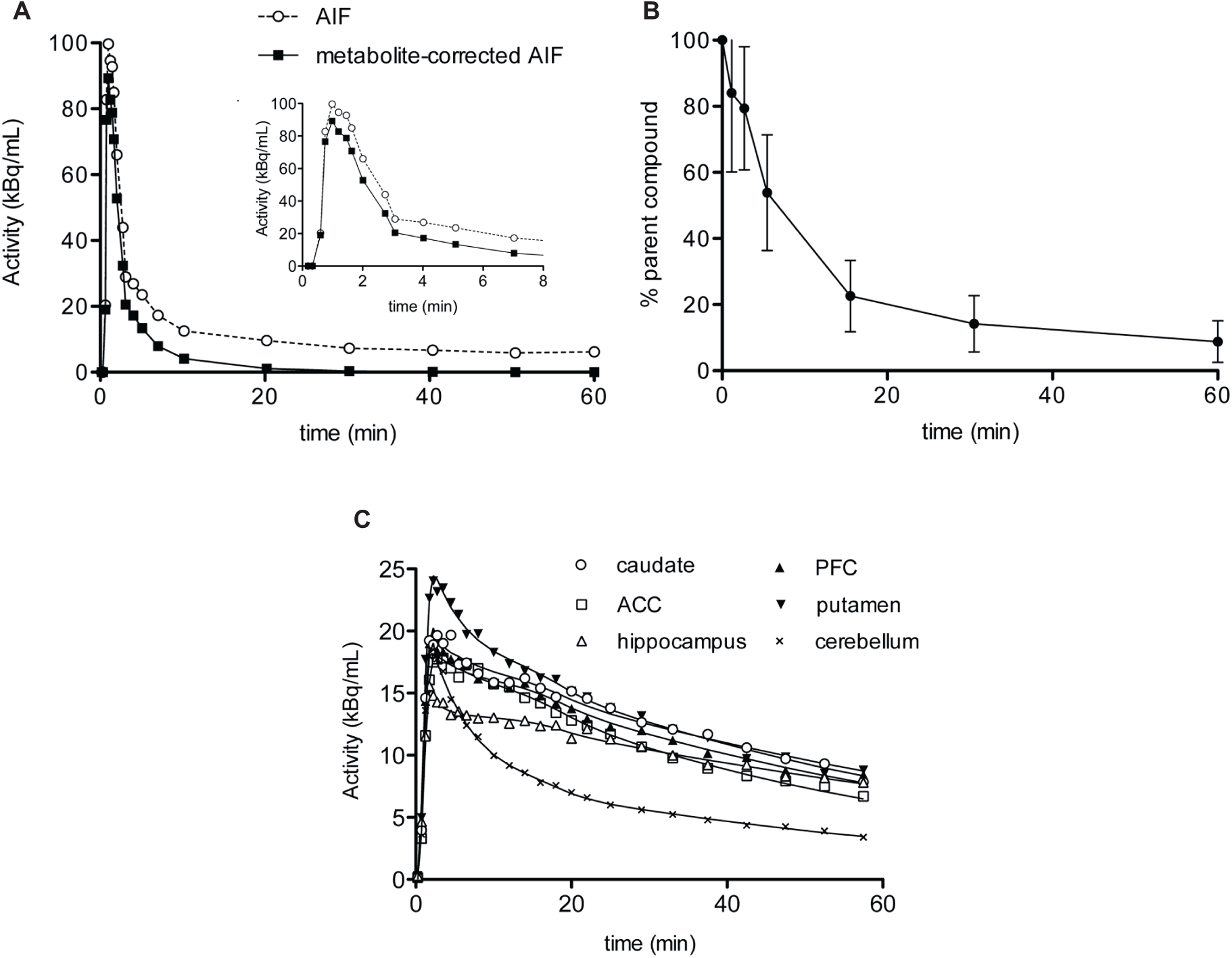
Plasma and regional time-activity curves for [^11^C]ABP688 and radiotracer metabolism. (**A**) Time-activity curves for the concentration of total radioactivity in plasma (dashed line) and of unchanged radioactivity in plasma (solid line) after correction for metabolites in a representative subject. AIF: arterial input function. The small insert shows the plasma peak in the first minutes. (**B**) Time course for the percentage of activity of parent compound in plasma (mean±S.D.; n = 6). (**C**) Representative regional time-activity curves after intravenous injection of [^11^C]ABP688 in a healthy subject. Symbols represent data for each brain region. Lines represent the fits to the data points with the unconstrained 2-tissue compartment model. ACC: anterior cingulate cortex; PFC: prefrontal cortex.

#### 3.2.2. Kinetic modeling using AIF

In the kinetic analysis, the 2TCM with unconstrained parameters adequately described all regional TACs. Representative fittings are depicted in Figure 3C. Unconstrained 2TCM yielded consistently lower AIC values than those obtained by fixing K_1_/k_2_ and was therefore preferred for all subsequent analysis (values not shown). Table 1 summarizes V_T_ values derived from the 2TCM analysis. These values were estimated with high precision, as attested by a mean percent of standard error of 8.5 ± 1.8% across subjects and brain regions. The V_T_ was highest in the PFC and ACC (3.7 ± 0.7 and 3.6 ± 0.6 mL/cm^3^ respectively) and lowest in the cerebellum (∼2.0 mL/cm^3^). No correlation between V_T_ and injected mass was found. There was an excellent agreement between the V_T_ values calculated from the 2TCM and those obtained using the GA approach (*r* = 0.99, R^2^ = 0.97, p<0.001; values in Table 1 and correlation graph with regression equation in Figure 4A).

**Figure 4.**
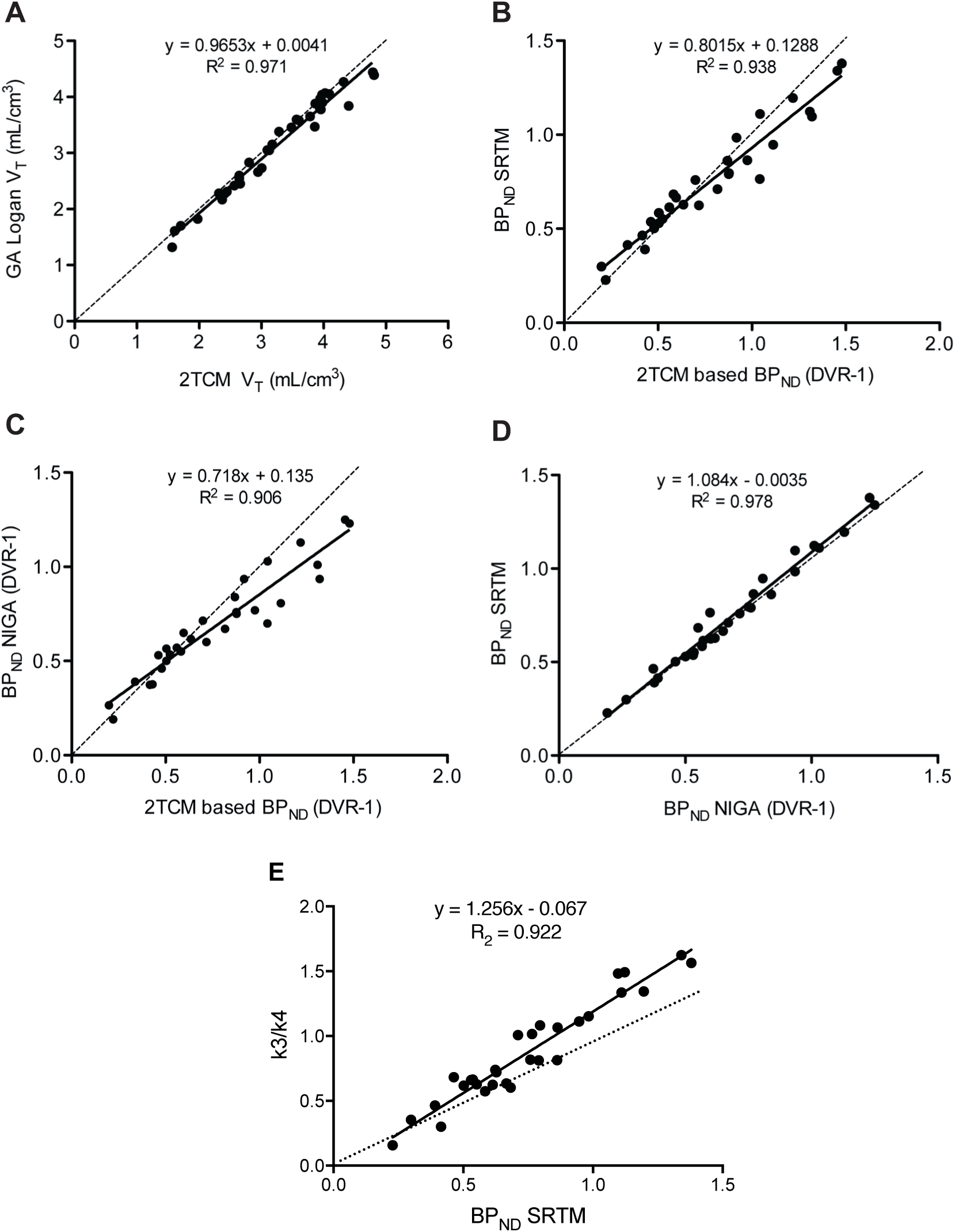
Correspondence of the outcome parameters distribution volume (V_T_) and binding potential (BP_ND_) derived by different methods of analysis. (**A**) Correlation between V_T_ values obtained by the 2-tissue compartment model (2TCM) and Logan’s graphical analysis (GA). (**B**) Correlation between BP_ND_ values derived from the V_T_ of 2TCM and the simplified reference tissue method (SRTM). (**C**) Correlation between BP_ND_ values obtained using the 2TCM and the distribution volume ratio (DVR-1) from Logan’s noninvasive GA (NIGA). (**D**) Correlation between BP_ND_ values obtained using the noninvasive reference tissue models SRTM and NIGA. (**E**) Correlation between SRTM-derived BP_ND_ and binding potentials directly derived from k3/k4 estimated with the constrained 2TCM. Data points for all brain regions from the 6 subjects are presented. Solid line represents linear regression analysis and dotted line represents identity. R^2^ derived from regression analysis are shown for each set, as well as regression equation.

The estimated rate constants are shown in Table 1. K_1_ mean values were relatively uniform across all brain regions, with highest values found in the putamen and lowest in the hippocampus. The range of K_1_ values (0.138–0.185 mL/cm^3^/min^-1^) correspond to a first-pass extraction of approximately 30%, considering reports of regional cerebral blood flow in humans. K_1_ standard error variability ranged from 4.4 to 8.8%. Larger, but tolerable, standard errors were found for k_2,_ k_3_ and k_4_ estimations, ranging from 10-23%, 11-24% and 8-19% respectively. Amongst the ROIs, the 2TCM fit provided consistently stronger identification of kinetic constants in the cerebellum and putamen. V_b_ had regional values ranging between 0.08 to 0.09 ml/cm^3^.

#### 3.2.3. Comparison of BP_ND_ measures in AIF and reference-based methods

BP_ND_ values derived from arterial input modeling were calculated from the V_T_ values of the target and reference regions in the 2TCM. Cerebellum was used as reference region in noninvasive quantifications. High correspondence between 2TCM BP_ND_ and the values derived from simplified noninvasive approaches was found (p<0.001; Figure 4B & 4C). In particular, SRTM provided the strongest agreement (r = 0.97, R^2^ = 0.94), although with a positive average bias of 6.4% in the putamen, and a negative bias of 7.8% in the hippocampus. As expected, noninvasive methods yielded lower intersubject variability compared to invasive model. NIGA approach provided BP_ND_ values in excellent correlation with SRTM (r = 0.99, R^2^ = 0.98, p<0.001), with comparable interindividual variability (36-40%) and a consistent underestimation of 5.0-11.0% (Figure 4D). The ratios of k3 over k4, with values derived from the constrained 2TCM, were in excellent agreement with BP_ND_ values from the SRTM (r = 0.96, R^2^ = 0.92, p<0.001) (Figure 4E). Mean BP_ND_ estimates across methods are summarized in Figure 5.

**Figure 5.**
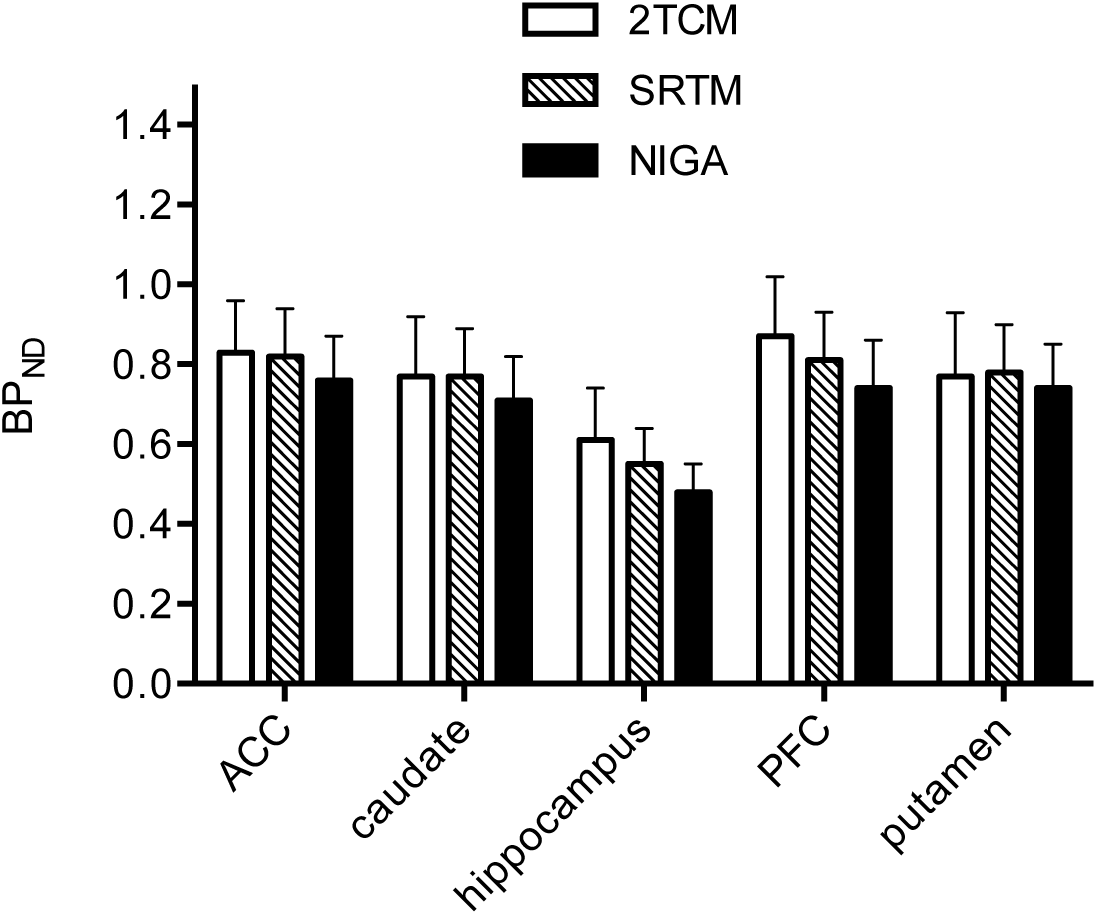
Regional binding potentials (BP_ND_) values estimated by three different methods of analysis: metabolite-corrected plasma input 2-tissue compartment model (2TCM); simplified reference tissue method (SRTM); noninvasive graphical analysis (NIGA). ACC: anterior cingulate cortex; PFC: prefrontal cortex. Data are presented as mean±S.E.M.

## 4. Discussion

Our study provides two major findings supporting the use of the cerebellum as a reference tissue for deriving [^11^C]ABP688 BP_ND_ values in humans. First, the availability of binding sites for [^3^H]ABP688 in the cerebellum was 15-fold lower than in the hippocampus. Second, there was a high correspondence between V_T_ or DVR estimated via 2TCM and BP_ND_ values using cerebellum as a reference region.

### 4.1. *In vitro* experiments

To our knowledge, this is the first study to investigate binding parameters in human cerebellum with [^3^H]ABP688 *in vitro* autoradiography and immunohistochemistry. Since the introduction of molecular agents for mGluR5, cerebellum has been described as a low uptake region when compared to regions such as the hippocampus (Hamill et al., 2005; Hintermann et al., 2007; Wyss et al., 2007; Helmenhorst et al., 2010). Consistent with these reports, we confirmed here the presence of small magnitude cerebellar specific binding in human subjects. The cerebellar concentration reported in our study (B_max_ of 0.08 pmol/mg tissue) is in agreement with our previous report for rat with the same techniques (B_max_ ∼ 0.09 pmol/mg tissue, with 15-fold hippocampus/cerebellum ratio) (Helmenhorst et al., 2010)

Autoradiography images (Figure 1) showed that the binding signal before and after competitor MPEP is similar to the background activity, supporting low cerebellar specific binding. In our *in vitro* experimental conditions, a 10 µmol/L blocking dose of MPEP imposed 95% [^3^H]ABP688 binding reduction in the hippocampus, and 75% in the cerebellum. In addition to a much smaller B_max_ compared to the hippocampus, the cerebellum shows a much lower ratio of specific to nonspecific binding. Moreover, the lower *K_D_* observed on the cerebellum suggests receptor’s affinity state is different from that in the hippocampus. It is improbable that the cerebellar specific binding sites reported here represents cross-binding (i.e., binding to mGluR1, another group I mGluR that is abundant in the cerebellum) owing to the high ABP688 specificity to mGluR5 (Hintermann et al., 2007). The low magnitude of [^3^H]ABP688 saturable binding sites as reported here agrees with earlier gene expression studies showing modest mGluR5 mRNA expression in the human adult cerebellum (Daggett et al., 1995; Berthele et al., 1999). At least, in the case of [^3^H]ABP688, the comparable mGluR5 binding parameters between rodents and humans support the assumption that use of cerebellum as reference region in the rat can be extended to humans.

Our [^3^H]ABP688 quantitative autoradiography results contrast with previous reports obtained with other mGluR5 allosteric ligands such as [^18^F]FPEB or [^11^C]MTEP, for which the brain/cerebellum ratio was determined as a 5-fold index. These discrepancies can be attributed to primate species evaluated and methodological differences (intact vs tissue homogenates), and/or method of quantification (quantitative autoradiography vs “non-wash wipe” assay”) (Patel et al., 2007). In addition, the reduced brain/cerebellum ratios previously reported for mGluR5 allosteric ligands might be in part secondary to the intrinsic pharmacokinetics properties of a given molecular probe (i.e., binding sites are not necessarily the same) (Patel et al., 2007; Hamill et al., 2005). Finally, cis/trans isomerism may also play a role on the quantification of binding parameters of different mGluR5 allosteric ligands such as [^3^H]ABP688, [^18^F]FPEB or [^11^C]MTEP. In our present study, we used a 1:8 cis/trans concentration of desmethyl-ABP688, which minimized the effects of the inactive cis-isomers (Ametamey et al., 2006). To the extent that these effects were present, they would have affected absolute binding values without altering correlations between values derived using different approaches.

Interestingly, when compared to the hippocampus, we found only a 26% mGluR5 reduction on cerebellar immunoreactivity in the presence of 15-fold reduction in the cerebellar [^3^H]ABP688 B_max_. These results are in agreement with a previous semi-quantitative study comparing mGluR5 immunoreactivity and allosteric binding site density (Patel et al., 2007) with the observation that immunohistochemistry is not a direct measurement for protein quantification. Although mGluR5 receptors are present in the cerebellum, the availability of their allosteric binding sites in comparison to mGluR5 located in the cortex and the hippocampus was markedly reduced. The biological interpretation of this dissociation is discussed below.

### 4.2. In vivo experiments

TAC analysis showed a rapid brain uptake with similar levels of initial radioactivity in all ROIs and cerebellum. Consistent with previous human studies using [^11^C]ABP688 (Ametamey et al., 2007; Treyer et al., 2007), activity in the cerebellum decreased more rapidly than elsewhere with similar trends over time, generating TACs with robust reproducibility across studies.

Similar to our previous work in rodents, here we directly compared AIF-based models to simplified reference tissue and graphical methods using the grey matter of the cerebellum as reference tissue. In the AIF-based method, the 2TCM configuration with four unconstrained parameters fitted the data from all regions, cerebellum included. This was expected, as previously shown for [^11^C]ABP688 in rats and humans (Treyer et al., 2007; Helmenhorst et al., 2010).

K_1_ values estimated with the 2TCM analysis showed excellent consistency within regions and between regions (ANOVA two-way, F = 1.59, p = 0.192), indicating uniform tracer delivery in the brain. V_T_ values in our study were similar to those reported from previous studies using an HRRT scanner (DeLorenzo et al., 2015 & 2017).

Interestingly, in line with reports from other tracers (Ginovart et al., 2007), a one-tissue compartment model does not adequately fit the cerebellum’s TAC. It is unlikely that this reflects the presence of a two-tissue compartment since the actual specific binding was shown to be minimal. More plausibly, this is due to a non-instantaneous equilibrium between free and nonspecific compartments that is usually assumed in the model simplification (Lammertsma et al., 1996b). The model choice for the cerebellum is further supported by the significant correspondence between V_T_ 2TCM-derived and V_T_ obtained with the Logan GA (Figure 4A), which does not consider compartments. The slight underestimation of GA V_T_ estimates (between 2 and 4%, depending on region) is common to many different tracers and likely due to statistical noise. Also, the possible contribution to a second compartment of a small (∼5%) fraction of radioactivity due to radiolabeled metabolites that entered the brain (Ametamey et al., 2006) cannot be excluded.

The ratio of k3 over k4 provides a direct estimate of the binding potential that is independent from the reference region. These values, when obtained with the constrained 2TCM, were highly correlated with the SRTM-derived BP_ND_ (Figure 4E), providing strong evidence that the cerebellum is a suitable reference region for mGluR5 quantification.

The data range for BP_ND_ was in line with other reports in baboons and humans (DeLorenzo et al., 2011a & 2011b; Akkus et al., 2013; Akkus et al., 2016). Despite the high correspondence between BP_ND_ estimates across the methods used, some quantitative differences were present (Figure 5). SRTM did not consistently underestimate BP_ND_, as the possible presence of a second compartment would have implied (Parsey et al., 2000). However, all averaged biases with respect to 2TCM estimates were in the acceptable limit of 7%. In support of the reliability of this non-invasive method, SRTM values were also robustly correlated with those obtained by NIGA. NIGA provided the lowest binding values among all methods, independently from ROI, and a stable weak underestimation, with a pinnacle for the smallest region hippocampus (-11%), with respect to SRTM. The latter should then be considered the most valid method for measuring occupancy in future group comparison studies.

When applying reference tissue models, it is assumed that V_ND_ is homogeneous in all brain regions and equal to the reference region V_T_. Since, as shown with autoradiography, specific binding in the cerebellum is negligible, only free and nonspecifically bound radiotracer significantly contributes to the cerebellum V_T_. In this study, cerebellum V_T_ was approximately 50-60% of that measured in mGluR5 -rich regions.

Arterial cannulation and blood sampling are invasive procedures that require additional effort and human resources. They impose discomfort to the tested individuals and the procedure is poorly accepted in certain clinical populations. In addition, the extraction of the metabolite-corrected AIF is one of the main sources of inter-subject variability. Therefore, non-invasive models for mGluR5 quantification with [^11^C]ABP688 using the cerebellum as reference region are preferable and reliable.

Any condition that might have an effect on nonspecific binding measures in the cerebellum potentially invalidates the initial assumption. In particular, cerebellar atrophy can compromise the accuracy of tracer level estimations because of partial volume effects (Lammertsma et al., 1996b). The sources of injury to the cerebellum varies, involving toxins (alcohol, chemotherapy, anticonvulsants), inflammation (autoantibodies, encephalitis), structural damage (stroke, tumors), inherited degenerations and congenital malformations.

### 4.3. Biological Interpretation and limitations of [^11^C]ABP688 BP_ND_

Although there is a good agreement between metabolite-corrected plasma input function and cerebellar reference tissue methods, the present results raise some caution on the interpretation of binding parameters involving molecular probes for mGluR5 allosteric binding sites. The 15-fold hippocampus/cerebellum ratio obtained with autoradiography contrasts with the 26% difference obtained with immunohistochemistry. Although both techniques serve to quantify mGluR5, the former technique relies on the availability of the mGluR5 allosteric binding site (tertiary conformation) and the latter on a sequence of amino acids (primary conformation) (Changeux & Edelsteinm, 2005). This dissociation suggests that mGluR5 allosteric binding sites are unavailable in cerebellum (Figure 6).

**Figure 6.**
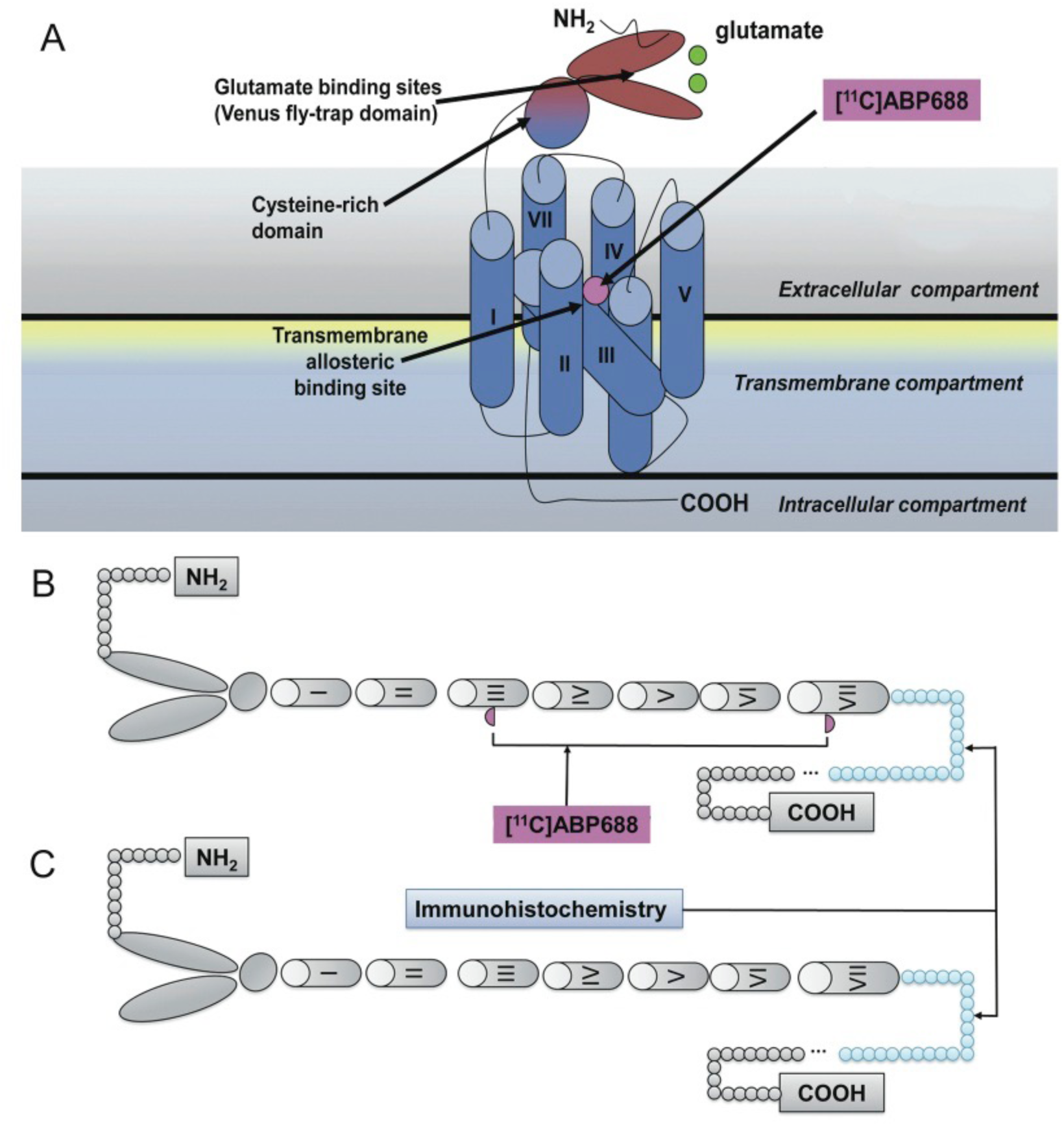
Top panel (**A**) represents mGluR5, a member of class C family of G-protein coupled receptors displaying a large extracellular amino-terminal domain for glutamate binding (dark blue), 7 transmembrane helical segments (light blue), and an allosteric pocket (violet). The lower panel shows a schematic representation of mGluR5 with (**B**) or without allosteric binding sites available for [^11^C]ABP688 binding (**C**). The former predominates in the prosencephalon and the latter in the cerebellum. The part painted in blue refers to the amino acid sequences (AA 1189-1212) recognized by the mGluR5 polyclonal antibody used in the study.

It has been proposed that the mGluR5 allosteric binding site is fundamentally a network of aromatic residues present between the 3 and 6 transmembrane loops. This binding site is highly dependent on the tertiary and quaternary receptor conformation (Changeux & Edelsteinm, 2005). In contrast, the mGluR5 antibody used in this study binds to a 21 amino acid sequence located in the mGluR5 C-terminus after protein denaturation and is therefore unaffected by receptor conformation changes (Malherbe et al., 2006). Thus, reduced binding sites due to either changes in primary or tertiary mGluR5 conformation might explain these results. Possibly, [^11^C]ABP688 binding captures a specific mGluR5 population abundant in the prosencephalon and scarce in the cerebellum. Our current data showing that [^11^C]ABP688 K_D_ in the cerebellum is about 4-fold lower than in the hippocampus (0.4 vs 1.5 nmol/L) implies that cerebellar receptors are in a higher affinity state. Interestingly, the mGluR5 splice variant most represented in the cerebellum is less sensitive to agonist-induced desensitization, with respect to other variants mostly expressed in the cerebral cortex and hippocampus (Malherbe et al., 2002).

These observations might also be explained by distinct cerebellar mGluR5 conformational states. Indeed, modifications in the receptor-radioligand interactions due to changes in affinity state has been described for D2 receptors, with D2 agonist PET ligands showing 2-fold displacement after amphetamine challenge compared to D2 antagonist ligands (Narendran et al., 2004). Glutamate release might reduce [^11^C]ABP688 affinity (Miyake et al., 2011) as its binding to the orthosteric site mobilizes the venus fly trap domain in the receptor’s N-terminus down towards the membrane, which could alter the accessibility to the allosteric binding sites residues in the transmembrane domains and thus affect [^11^C]ABP688 affinity. However, this claim must be confirmed by further studies in which autoradiography and receptor immunodetection could be compared head to head in healthy and disease conditions.

### 4.4. Limitations of the present study

The external validity of the present findings requires the consideration of the following limitations. For our *in vitro* experiments, systematic comparisons between mGluR5 immunohistochemistry and binding parameters were not conducted since tissue sections used in the two techniques were derived from different sets of brains (frozen or formalin fixed). However, it is important to emphasize that there was no age or ante mortem clinical difference between the two subject populations from which the specimens were derived. Together, our *in vitro* experiments suggest that cerebellar mGluR5 differ qualitatively and quantitatively between the cerebellum and prosencephalon. For the purpose of the present study, these data support the use of the cerebellum as reference tissue in [^11^C]ABP688 binding quantification.

Our *in vivo* experiments involved a relatively small group of healthy volunteers. In addition, it would be highly desirable to conduct these experiments with injections of the active trans- [^11^C]ABP688, although the radiochemistry used in this study results in a ratio of 1:8 (Ametamey et al., 2006). Nevertheless, the isomer related effect would only change the absolute V_T_/BP_ND_ values quantification without affecting the ability to examine correlations between values calculated with different methods. The use of the HRRT scanner might bias our results due to its higher sensitivity and known issues regarding scatter and attenuation correction at the level of cerebellum. Since comparison across multiple kinetic models were beyond the scope of the present manuscript, the kinetic analyses presented here focuses on widely available methodology.

In summary, our data on reproducibility obtained in rats showed favorable results. Overall, there was excellent agreement between estimates of BP_ND_ and metabolite-corrected plasma derived values, demonstrating the validity of models relying on reference region to quantify mGlu5 receptor availability with [^11^C]ABP688.

## Disclosure/Conflict of Interest

The authors declare no conflict of interest. The views expressed herein do not necessarily represent the views of the Canadian Minister of Health or the Government of Canada.

## Data availability

All software and procedures concerning the data acquisition and analysis have been detailed in the Methods section. All data sets are available from the corresponding author upon reasonable request.

## Author contributions

Michele S Milella: Investigation, Formal analysis, Data curation, Methodology, Visualization, and Writing - original draft. Luciano Minuzzi: Investigation, and Data curation. Chawki Benkelfat: Supervision, and Writing - review & editing. Jean P Soucy: Resources, Supervision, and Writing - review & editing. Alexandre Kirlow: Resources. Esther Schirrmacher: Resources, and Supervision. Mark Angle: Resources, and Supervision. Jeroen AJ Verhaeghe: Writing - review & editing. Gassan Massarweh: Resources, and Supervision. Andrew J Reader: Supervision, and Writing - review & editing. Antonio Aliaga: Investigation, and Methodology. Jose Eduardo Peixoto-Santos: Investigation, and Data curation. Marie-Christine Guiot: Resources, and Supervision. Eliane Kobayashi: Writing - review & editing. Pedro Rosa-Neto: Conceptualization, Funding Aquisition, Resources, Supervision, Data curation, Software, Methodology, and Writing - review & editing. Marco Leyton: Conceptualization, Funding Aquisition, Resources, Supervision, Data curation, and Writing - review & editing.

## Acknowledgements

We extend our thanks to the PET Radiochemistry team at Montreal Neurological Institute for providing us access to the tracer and to the PET scanner. This work was supported in part by Alzheimer’s Association [NIRG-08-92090 to PR-N], Nussia & André Aisenstadt Foundation [PR-N], Fonds de la recherche en santé du Québec [16326, PR-N], and the Canadian Institutes of Health Research [MOP-115131 to PR-N and MOP-36429 to ML]. The study benefited from the financial support of Health Canada, through the Canada Brain Research Fund, an innovative partnership between the Government of Canada (through Health Canada) and Brain Canada, and the Montreal Neurological Institute.

## Notes

### Competing Interest Statement

The authors have declared no competing interest.

### Funding Statement

This work was supported in part by Alzheimer's Association [NIRG-08-92090 to PR-N], Nussia & Andre Aisenstadt Foundation [PR-N], Fonds de la recherche en sante du Quebec [16326, PR-N], and the Canadian Institutes of Health Research [MOP-115131 to PR-N and MOP-36429 to ML]. The study benefited from the financial support of Health Canada, through the Canada Brain Research Fund, an innovative partnership between the Government of Canada (through Health Canada) and Brain Canada, and the Montreal Neurological Institute.

### Author Declarations

All in vitro experiments were carried out in accordance with the guidelines provided by the Douglas Brain Bank research board, approved by the Research and Ethics Board of the Douglas Research Institute, McGill University. The in vivo study was carried out in accordance with the Declaration of Helsinki and was approved by the Research and Ethics Board of the Montreal Neurological Institute/McGill University. Written informed consent with details of the experimental procedures and approved by our institution Research Ethics Board was obtained from all subjects before scan sessions

## References

Akkus, F., Ametamey, S.M., Treyer, V., Burger, C., Johayem, A., Umbricht, D., Gomez Mancilla, B., Sovago, J., Buck, A., & Hasler, G. (2013). Marked global reduction in mGluR5 receptor binding in smokers and ex-smokers determined by [11C] ABP688 positron emission tomography. PNAS, 110(2), 737-742. 10.1073/pnas.1210984110

Akkus, F., Treyer, V., Johayem, A., Ametamey, S.M., Mancilla, B.G., Sovago, J., Buck, A., & Hasler, G. (2016). Association of Long-Term Nicotine Abstinence With Normal Metabotropic Glutamate Receptor-5 Binding. Biol Psychiatry 15;79(6):474-80. 10.1016/j.biopsych.2015.02.027

Ametamey, S.M., Kessler, L.J., Honer, M., Wyss, M.T., Buck, A., Hintermann, S., Auberson, Y.P., Gasparini, F., & Schubiger, P.A. (2006). Radiosynthesis and preclinical evaluation of 11C-ABP688 as a probe for imaging the metabotropic glutamate receptor subtype 5. J Nuclear Medicine, 47, 698-705.

Ametamey, S.M., Treyer, V., Streffer, J., Wyss, M.T., Schmidt, M., Blagoev, M., Hintermann, S., Auberson, Y., Gasparini, F., Fischer, U.C., & Buck, A. (2007). Human PET studies of metabotropic glutamate receptor subtype 5 with 11C-ABP688. J Nuclear Medicine, 48, 247-252

Berthele, A., Platzer, S., Laurie, D.J., Weis, S., Sommer, B., Zieglgänsberger, W., Conrad, B., & Tölle, T.R. (1999). Expression of metabotropic glutamate receptor subtype mRNA (mGluR1-8) in human cerebellum. Neuroreport, 10, 3861-3867. 10.1097/00001756-199912160-00026

Breysse, N., Baunez, C., Spooren, W., Gasparini, F., & Amalric, M. (2002). Chronic but not acute treatment with a metabotropic glutamate 5 receptor antagonist reverses the akinetic deficits in a rat model of parkinsonism. Journal of neuroscience, 22,5669-5678. 10.1523/JNEUROSCI.22-13-05669.2002

Bruno, V., Battaglia, G., Copani, A., D’Onofrio, M., Di Iorio, P., De Blasi, A., Melchiorri, D., Flor, P.J., & Nicoletti, F. (2001). Metabotropic glutamate receptor subtypes as targets for neuroprotective drugs. J Cereb Blood Flow Metab, 21, 1013-1033. 10.1097/00004647-200109000-00001

Changeux, J-P., & Edelsteinm, S.J. (2005). Allosteric mechanisms of signal transduction. Science, 308, 1424-1428. 10.1126/science.1108595

Chiamulera, C., Epping-Jordan, M.P., Zocchi, A., et al. (2001). Reinforcing and locomotor stimulant effects of cocaine are absent in mGluR5 null mutant mice. Nat neurosci, 4, 873-874. 10.1038/nn0901-873

Costes, N., Dagher, A., Larcher, K., Evans, A.C., Collins, D.L., & Reilhac, A. (2009). Motion correction of multi-frame PET data in neuroreceptor mapping: simulation based validation. NeuroImage, 47, 1496-1505. 10.1016/j.neuroimage.2009.05.052

Cox, S.M., Tippler, M., Jaworska, N., Smart K, Castellanos-Ryan, N., Durand, F., Allard, D., Benkelfat, C., Parent, S., Dagher, A., Vitaro, F., Boivin, M., Pihl, R.O., Côté, S., Tremblay, R.E., Séguin, J.R., & Leyton, M. (2020). mGlu5 Receptor availability in youth at risk for addictions: Effects of vulnerability traits and cannabis use. Neuropsychopharmacology, 45(11), 1817-1825. 10.1038/s41386-020-0708-x

Daggett, L.P., Sacaan, A.I., Akong, M., Rao, S.P., Hess, S.D., Liaw, C., Urrutia, A., Jachec, C., Ellis, S.B., Dreessen, J., et al. (1995). Molecular and functional characterization of recombinant human metabotropic glutamate receptor subtype 5. Neuropharmacology, 34, 871-886. 10.1016/0028-3908(95)00085-k

de Jong, H.W., van Velden, F.H., Kloet, R.W., Buijs, F.L., Boellaard. R., & Lammertsma, A.A. (2007). Performance evaluation of the ECAT HRRT: an LSO-LYSO double layer high resolution, high sensitivity scanner. Phys med biol, 52, 1505-1526. 10.1088/0031-9155/52/5/019

DeLorenzo, C., Kumar, J.S., Mann, J.J., & Parsey, R.V. (2011a). In vivo variation in metabotropic glutamate receptor subtype 5 binding using positron emission tomography and [11C]ABP688. J Cereb Blood Flow Metab, 31, 2169-2180. 10.1038/jcbfm.2011.105

DeLorenzo, C., Milak, M.S., Brennan, K.G., Kumar, J.S., Mann, J.J., & Parsey, R.V. (2011b). In vivo positron emission tomography imaging with [11C]ABP688: binding variability and specificity for the metabotropic glutamate receptor subtype 5 in baboons. European journal of nuclear medicine and molecular imaging, 38, 1083-1094. 10.1007/s00259-010-1723-7

DeLorenzo, C., DellaGioia, M., Bloch, M., Sanacora, G., Nabulsi, N., Abdallah, C., Yang, J., Wen, R., Mann, J.J., Krystal, J.H., Parsey, R.V., Carson, R.E., & Esterlis, I. (2015). In vivo ketamine-induced changes in [11C]ABP688 binding to metabotropic glutamate receptor subtype 5. Biol Psych, 77(3), 266-275. 10.1016/j.biopsych.2014.06.024

DeLorenzo, C., Gallezot, J.D., Gardus, J., Yang, J., Planeta, B., Nabulsi, N., Ogden, R.T., Labaree, D.C., Huang, Y.H., Mann, J.J., Gasparini, F., Lin, X., Javitch, J.A., Parsey, R.V., Carson, R.E., & Esterlis, I. (2017). In vivo variation in same-day estimates of metabotropic glutamate receptor subtype 5 binding using [11C]ABP688 and [18F]FPEB. J Cereb Blood Flow Metab, 37(8), 2716-2727. 10.1177/0271678X16673646

Elmenhorst, D., Minuzzi, L., Aliaga, A., Rowley, J., Massarweh, G., Diksic, M, Bauer, A., & Rosa-Neto, P. (2010). In vivo and in vitro validation of reference tissue models for the mGluR(5) ligand [(11)C]ABP688. J Cereb Blood Flow Metab, 30, 1538-1549. 10.1038/jcbfm.2010.65

Gasparini, F., Bilbe, G., Gomez-Mancilla, B., & Spooren, W. (2008). mGluR5 antagonists: discovery, characterization and drug development. Current opinion in drug discov & development, 11, 655-665

Ginovart, N., Willeit, M., Rusjan, P., Graff, A., Bloomfield, P.M., Houle, S., Kapur, S., & Wilson, A.A. (2007). Positron emission tomography quantification of [11C]-(+)-PHNO binding in the human brain. J Cerebral Blood Flow Metab, 27, 857-871. 10.1038/sj.jcbfm.9600411

Gladding, C.M., Fitzjohn, S.M., & Molnar, E. (2009). Metabotropic glutamate receptor-mediated long-term depression: molecular mechanisms. Pharmacol rev, 61, 395-412. 10.1124/pr.109.001735

Hamill, T.G., Krause, S., Ryan, C., Bonnefous, C., Govek, S., Seiders, T.J., Cosford, N.D., Roppe, J., Kamenecka, T., Patel, S., Gibson, R.E., Sanabria, S., Riffel, K., Eng, W., King, C., Yang, X., Green, M.D., O’Malley, S.S., Hargreaves, R., & Burns, H.D. (2005). Synthesis, characterization, and first successful monkey imaging studies of metabotropic glutamate receptor subtype 5 (mGluR5) PET radiotracers. Synapse, 56, 205-216. 10.1002/syn.20147

Hintermann, S., Vranesic, I., Allgeier, H., Brülisauer, A., Hoyer, D., Lemaire, M., Moenius, T., Urwyler, S., Whitebread, S., Gasparini, F., & Auberson, Y.P. (2007). ABP688, a novel selective and high affinity ligand for the labeling of mGlu5 receptors: identification, in vitro pharmacology, pharmacokinetic and biodistribution studies. Bioorganic & medicinal chemistry, 15, 903-914. 10.1016/j.bmc.2006.10.038

Innis, R.B., Cunningham, V.J., Delforge, J., Fujita, M., Gjedde, A., Gunn, R.N., Holden, J., Houle, S., Huang, S.C., Ichise, M., Iida, H., Ito, H., Kimura, Y., Koeppe, R.A., Knudsen, G.M., Knuuti, J., Lammertsma, A.A., Laruelle, M., Logan, J., Maguire, R.P., Mintun, M.A., Morris, E.D., Parsey, R., Price, J.C., Slifstein, M., Sossi, V., Suhara, T., Votaw, J.R., Wong, D.F., & Carson, R.E. (2007). Consensus nomenclature for in vivo imaging of reversibly binding radioligands. J Cereb Blood Flow Metab, 27, 1533-1539. 10.1038/sj.jcbfm.9600493

Kew, J.N.C., & Kemp, J.A. (2005). Ionotropic and metabotropic glutamate receptor structure and pharmacology. Psychopharmacology, 179, 4-29. 10.1007/s00213-005-2200-z

Kjærgaard, K., Frisch, K., Sørensen, M., Munk, O.L., Hofmann, A.F., Horsager, J., Schacht, A.C., Erickson, M., Shapiro, D., & Keiding, S. (2021). Obeticholic acid improves hepatic bile acid excretion in patients with primary biliary cholangitis. J Hepatol, 74(1), 58-65. 10.1016/j.jhep.2020.07.028

Lammertsma, A.A., & Hume, S.P. (1996). Simplified reference tissue model for PET receptor studies. NeuroImage, 4, 153-158. 10.1006/nimg.1996.0066

Lammertsma, A.A., Bench, C.J., Hume, S.P., Osman, S., Gunn, K., Brooks, D.J., & Frackowiak, R.S. (1996b). Comparison of methods for analysis of clinical [11C]raclopride studies. J Cerebral Blood Flow Metab, 16, 42-52. 10.1097/00004647-199601000-00005

Liberatore, G.T., Wong, J.Y., Krenus, D., Jeffreys, B.J., Porritt, M.J., & Howells, D.W. (1999). Tissue fixation prevents contamination of tritium-sensitive storage phosphor imaging plates. Biotechniques, 26, 432-434. 10.2144/99263bm13

Logan, J., Fowler, J.S., Volkow, N.D., Wolf, A.P., Dewey, S.L., Schlyer, D.J., MacGregor, R.R., Hitzemann, R., Bendriem, B., Gatley, S.J., et al. (1990). Graphical analysis of reversible radioligand binding from time-activity measurements applied to [N-11C-methyl]-(-)-cocaine PET studies in human subjects. J Cereb Blood Flow Metab, 10, 740-747. 10.1038/jcbfm.1990.127

Logan, J., Fowler, J.S., Volkow, N.D., Wang, G.J., Ding, Y.S., & Alexoff, D.L. (1996). Distribution volume ratios without blood sampling from graphical analysis of PET data. J J Cereb Blood Flow Metab, 16, 834-840. 10.1097/00004647-199609000-00008

Malherbe, P., Kew, J.N., Richards, J.G., Knoflach, F., Kratzeisen, C., Zenner, M.T., Faull, R.L., Kemp, J.A., & Mutel, V. (2002). Identification and characterization of a novel splice variant of the metabotropic glutamate receptor 5 gene in human hippocampus and cerebellum. Brain res Mol brain res, 109, 168-178. 10.1016/s0169-328x(02)00557-0

Malherbe, P., Kratochwil, N., Muhlemann, A., Zenner, M.T., Fischer, C., Stahl, M., Gerber, P.R., Jaeschke, G., & Porter, R.H. (2006). Comparison of the binding pockets of two chemically unrelated allosteric antagonists of the mGlu5 receptor and identification of crucial residues involved in the inverse agonism of MPEP. J neurochem, 98, 601-615. 10.1111/j.1471-4159.2006.03886.x

Martinez, D., Slifstein, M., Nabulsi, N., Grassetti, A., Urban, N.B., Perez, A., Liu, F., Lin, S.F., Ropchan, J., Mao, X., Kegeles, L.S., Shungu, D.C., Carson, R.E., & Huang, Y. (2014). Imaging glutamate homeostasis in cocaine addiction with the metabotropic glutamate receptor 5 positron emission tomography radiotracer [(11)C]ABP688 and magnetic resonance spectroscopy. Biol Psychiatry, 75(2), 165-71. 10.1016/j.biopsych.2013.06.026

Milella, M.S., Marengo, L., Larcher, K., Fotros, A., Dagher, A., Rosa-Neto, P., Benkelfat, C., & Leyton, M. (2014). Limbic system mGluR5 availability in cocaine dependent subjects: a high-resolution PET [(11)C]ABP688 study. Neuroimage, 98, 195-202. 10.1016/j.neuroimage.2014.04.061

Miyake, N., Skinbjerg, M., Easwaramoorthy, B., Kumar, D., Girgis, R.R., Xu, X., Slifstein, M., & Abi-Dargham, A. (2011). Imaging changes in glutamate transmission in vivo with the metabotropic glutamate receptor 5 tracer [11C] ABP688 and N-acetylcysteine challenge. Biol psychiatry, 69, 822-824. 10.1016/j.biopsych.2010.12.023

Narendran, R., Hwang, D.R., Slifstein, M., Talbot, P.S., Erritzoe, D., Huang, Y., Cooper, T.B., Martinez, D., Kegeles, L.S., Abi-Dargham, A., & Laruelle, M. (2004). In vivo vulnerability to competition by endogenous dopamine: comparison of the D2 receptor agonist radiotracer (-)-N-[11C]propyl-norapomorphine ([11C]NPA) with the D2 receptor antagonist radiotracer [11C]-raclopride. Synapse, 52, 188-208. 10.1002/syn.20013

Parameshwaran, K., Dhanasekaran, M., & Suppiramaniam, V. (2008). Amyloid beta peptides and glutamatergic synaptic dysregulation. Experimental neurology, 210, 7-13. 10.1016/j.expneurol.2007.10.008

Parsey, R.V., Slifstein, M., Hwang, D.R., Abi-Dargham, A., Simpson, N., Mawlawi, O., Guo, N.N., Van Heertum, R., Mann, J.J., & Laruelle, M. (2000). Validation and reproducibility of measurement of 5-HT1A receptor parameters with [carbonyl-11C]WAY-100635 in humans: comparison of arterial and reference tisssue input functions. J Cereb Blood Flow Metab, 20, 1111-1133. 10.1097/00004647-200007000-00011

Patel, S., Hamill, T.G., Connolly, B., Jagoda, E., Li, W., & Gibson, R.E. (2007). Species differences in mGluR5 binding sites in mammalian central nervous system determined using in vitro binding with [18F]F-PEB. Nucl Med Biol, 34, 1009-1017. 10.1016/j.nucmedbio.2007.07.009

Reader, A.J., Sureau, F.C., Comtat, C., Trebossen, R., & Buvat, I. (2006). Joint estimation of dynamic PET images and temporal basis functions using fully 4D ML-EM. Physics in medicine and biology, 51, 5455-5474. 10.1088/0031-9155/51/21/005

Shigemoto, R., Nomura, S., Ohishi, H., Sugihara, H., Nakanishi, S., & Mizuno, N. (1993). Immunohistochemical localization of a metabotropic glutamate receptor, mGluR5, in the rat brain. Neuroscience letters, 163, 53-57. 10.1016/0304-3940(93)90227-c

Spooren, W.P., Vassout, A., Neijt HC, Kuhn R, Gasparini, F., Roux, S., Porsolt, R.D., Gentsch, C. (2000). Anxiolytic-like effects of the prototypical metabotropic glutamate receptor 5 antagonist 2-methyl-6-(phenylethynyl)pyridine in rodents. The Journal of pharmacology and experimental therapeutics, 295, 1267-1275

Treyer, V., Streffer, J., Wyss, M.T., Bettio, A., Ametamey, S.M., Fischer, U., Schmidt, M., Gasparini, F., Hock, C., & Buck, A. (2007). Evaluation of the metabotropic glutamate receptor subtype 5 using PET and 11C-ABP688: assessment of methods. J nuclear medicine, 48, 1207-1215.

van Velden, F.H., Kloet, R.W., van Berckel, B.N., Buijs, F.L., Luurtsema, G., Lammertsma, A.A., & Boellaard, R. (2009). HRRT versus HR+ human brain PET studies: an interscanner test-retest study. J nuclear medicine, 50, 693-702. 10.2967/jnumed.108.058628

Wyss, M.T., Ametamey, S.M., Treyer, V., Bettio, A., Blagoev, M., Kessler, L.J., Burger, C., Weber, B., Schmidt, M., Gasparini, F., & Buck, A. (2007). Quantitative evaluation of 11C-ABP688 as PET ligand for the measurement of the metabotropic glutamate receptor subtype 5 using autoradiographic studies and a beta-scintillator. NeuroImage, 35, 1086-1092. 10.1016/j.neuroimage.2007.01.005

