## Supplementary information for "Quantification of [^11^C]ABP688 binding in human brain using cerebellum as reference region: biological interpretation and limitations"

**Table 1.** Demographics from donors of tissue specimens used in the present study. There were no group differences (AR vs. IHC) in age ($p=0.9$; $t=0.09$; $df=9$) or postmortem processing time to freezing/fixation ($p=0.2$; $t=1.38$; $df=9$).

| Specimen | Experiment | Age range | Sex | Postmortem Delay (hours) | Side | Cause of Death/Diagnosis |
|---|---|---|---|---|---|---|
| 1 | AR | 70-80 | M | 20.5 | R | Larynx neoplasia |
| 2 | AR | 80-90 | F | 32.5 | R | Acute Myocardial Infarct |
| 3 | AR | 70-80 | F | 19.7 | R | Renal failure |
| 4 | AR | 40-50 | F | 43.7 | L | Lung neoplasia |
| 5 | AR | 70-80 | F | 17.2 | L | Acute Myocardial Infarct |
| 6 | IHC | 90-100 | F | 23.75 | L | Hepatic Metastasis |
| 7 | IHC | 80-90 | M | 26.75 | L | Lung Carcinoid Tumor |
| 8 | IHC | 80-90 | M | 8 | L | Ruptured abdominal aortic aneurism |
| 9 | IHC | 50-60 | F | 26.25 | L | Metastatic lung cancer |
| 10 | IHC | 60-70 | M | 8.75 | L | Pancreatic neoplasm |
| 11 | IHC | 50-60 | M | 17.67 | L | Gastric neoplasm |

Legends: AR (autoradiography), IHC (immunohistochemistry), F (female), M (male), R (right), L (left).
